# Gait-Related Digital Mobility Outcomes in Parkinson’s Disease: New Insights into Convergent Validity?

**DOI:** 10.64898/2026.03.07.26347847

**Authors:** Cyrille E. Mvomo, Jordan Bedime Songue Njiki, Dahlia Leibovich, Clara Guedes, Alexandra Potvin-Desrochers, Philippe C. Dixon, Chris Easthope Awai, Caroline Paquette

## Abstract

**Objective:** In Parkinson’s disease (PD), gait-related digital mobility outcomes (DMOs) show promise for monitoring mobility decline, but convergent validity remains limited. To improve convergent validity, demonstrating convergence with motor severity scales and PD-specific neural mechanisms underlying mobility has been proposed. However, severity scales capture both PD-specific and non-specific factors. Requiring mechanistic evidence may favor DMOs that converge with underlying mechanisms, while those converging only with severity scales may be overlooked despite capturing broader mobility dimensions. Here, we asked whether PD-specific neural mechanisms underlying mobility enhanced the convergence of DMOs with motor severity scales – so that integrating mechanistic evidence in validation could improve (and not impede) convergent validity.

**Method:** Principal component analysis was applied to task-based functional neuroimaging data to identify a measure associated with PD motor network dysfunction. An optimization problem was then formulated in which deep learning examined the convergence of a signal-based DMO with motor severity in laboratory and real-world contexts, and Tracing Gradient Descent assessed the influence of the identified measure on DMO–severity convergence.

**Results:** Greater PD motor network dysfunction was associated with reduced Attractor Complexity Index (ACI) values (*ρ*=−0.54). Strong DMO–severity convergence was found across contexts (*ρ*=|0.81−0.82|). Reduced ACI (i.e., greater PD motor network dysfunction) markedly enhanced DMO–severity convergence across contexts (*r*_*rb*_=|0.63−0.29|).

**Conclusion:** PD-specific neural mechanisms underlying mobility enhanced the convergence of a DMO with severity scales.

**Significance:** Integrating mechanistic validation into DMO validation could improve convergent (and construct) validity, a prerequisite for regulatory approval and adoption in clinical trials and practice.

## I. Introduction

IN Parkinson’s disease (PD), progressive neurodegeneration across multiple neurotransmitter systems and medication-related complications produce motor symptoms that impair mobility [1]. As mobility worsens, independence and quality of life decline [1]. Preserving mobility is therefore a priority for people with PD (PwP) and their caregivers, and effective management requires close monitoring to guide treatment adjustments [1], [2]. Clinicians traditionally monitor mobility using rating scales such as the Movement Disorder Society Unified Parkinson’s Disease Rating Scale part III (MDS-UPDRS III) [3]. Although validated, these scales present limitations. They lack sensitivity to subtle yet meaningful mobility decline, require episodic in-person administration by trained specialists, and are semi-subjective [3]–[6]. Consequently, advances in wearable sensing have motivated the development of digital mobility outcomes (DMOs) to address the limitations of clinical scales [7]. Among them, gait-related DMOs represent the most mature class [8]–[10]. Using a minimal number of wearable sensors, gait DMOs can enable sensitive, objective, scalable, remote, reliable, continuous, and passive monitoring of mobility [8]–[10]. These characteristics position gait DMOs as promising tools for both clinical practice and research [8]–[10].

Despite this promise, the convergent validity (i.e., agreement with known measures of the same construct) of gait DMOs remains insufficiently established. A major obstacle lies in the absence of an optimal reference measure of mobility. Without such a reference, validation studies infer convergent validity primarily from convergence with the same limited clinical scales that DMOs aim to improve [11]–[13]. This circular approach raises doubts about the true convergent validity of gait DMOs [14], [15], particularly when they are evaluated in the real world, where multiple sources of variability blur the distinction between mobility-related and unrelated features [16]. Addressing this issue is essential because convergent validity constitutes a core component of construct validity, and demonstrating construct validity remains a prerequisite for regulatory approval and clinical adoption of gait DMOs [17].

Accordingly, recent proposals suggest moving toward a multilevel validation framework [14], [18], [19]. Under such a framework, examining the convergence of a candidate gait DMO with neural mechanisms specific to motor dysfunction in PD, in addition to its convergence with clinical scales, could strengthen its convergent validity [18]. Although this view is appealing, a conflicting view exists. Observational studies indicate that mobility decline in PwP arises not only from PD-specific factors but also from non-specific factors such as aging and comorbidities [20], [21]. Clinical scales capture both PD-specific and non-specific factors and therefore reflect broader aspects of mobility than those specific to PD alone [3], [21]. Under this view, requiring PD-specific mechanistic evidence in the validation process may disqualify a candidate gait DMO that shows no convergence with neural mechanisms of PD motor decline, despite demonstrating convergence with clinical scales and therefore capturing a broader representation of mobility. Resolving the conflict between these two views requires determining whether and how neural mechanisms specific to motor dysfunction in PD influence the convergence of gait DMOs with clinical scales. If PD-specific neural mechanisms of motor dysfunction enhance convergence with clinical scales, integrating mechanistic evidence alongside clinical evidence could only strengthen convergent (and thus construct) validity by: (i) improving the capture of PD-specific aspects of mobility through direct neural anchoring, while (ii) retaining broader non-specific aspects through clinical scales, thus preserving a broad representation of mobility in PwP.

Our preliminary findings suggested that a gait DMO can be sensitive to both clinical scales and PD-specific neural mechanisms of motor dysfunction [22]. However, whether neural mechanisms specific to motor dysfunction in PD enhance convergence with clinical scales remains unknown. Addressing this question first requires identifying a trait associated with underlying PD motor deterioration that can be quantified during gait. One candidate trait is loss of gait automaticity. Dysfunction of the striato-pallido-thalamo-cortico-cerebellar circuit, which constitutes the PD motor network [23], represents a hallmark neural mechanism of PD motor impairment and manifests during gait as reduced automaticity and flexibility, and increased reliance on attentional control [24], [25]. This loss of automaticity becomes more pronounced as the disease progresses [24]. Recent advances in nonlinear dynamics have introduced metrics such as the Attractor Complexity Index (ACI), which enable quantification of gait automaticity during walking tasks of graded complexity [26], [27]. Although ACI has shown promise, direct evidence supporting its relationship with PD motor network dysfunction remains absent. Therefore, this work expanded on our initial study and aimed to: (1) introduce ACI as a measure of PD motor network dysfunction and (2) use it as a proxy to examine whether and how PD motor network dysfunction influences the convergence of a gait DMO with clinical scales of motor severity in PwP (referred to as clinical convergence).

First, we characterized PD motor network dysfunction during gait by applying a principal component analysis (PCA)–based algorithm to task-based functional neuroimaging data acquired during straight and complex walking. Analyses revealed that reduced ACI is associated with greater PD motor network dysfunction. Second, ACI was employed in two case studies in which an optimization problem was formulated to examine its influence on the clinical convergence of a previously validated gait DMO. We re-examined the DMO in laboratory (case study 1) and home contexts (case study 2). Examining both contexts represents a key feature of this work because distinct constructs of mobility exist: mobility capacity, which reflects what an individual can do under scripted evaluation, and mobility performance, which reflects what an individual does in daily life [17]. A signal-based DMO reflecting movement magnitude during gait was selected [22], [28], [29]. This DMO was chosen because of its computational simplicity (i.e., minimal processing requirements that reduce bias and promote reproducibility) [16]. It was also selected because its convergence with clinical scales has previously been assessed using deep learning [22], [28], which enables modelling of non-monotonic relationships between digital and clinical measures that are characteristic of PD [30] but are missed by classical approaches (e.g., correlation). Furthermore, modern post-hoc explainable artificial intelligence (XAI) methods such as Tracing Gradient Descent (TracIn) now allow granular quantification of the mechanisms that influence such complex associations during optimization [31]. Deep learning was therefore applied and confirmed the strong clinical convergence of the DMO using larger and more diverse datasets than in our initial work [22]. TracIn then showed that lower ACI (i.e., greater PD motor network dysfunction) markedly enhances clinical convergence.

## II. Materials and Methods

### A. Quantifying PD Motor Network Dysfunction via AC

#### 1) Dataset

Data from 18 PwP (33% female; median ± IQR age: 65 ± 7 years) and 7 healthy controls (HC) (43% female; median ± IQR age: 59 ± 2 years) were analyzed. These data were collected in two observational studies designed to measure gait capacity and brain function [32], [33], and together constitute the *laboratory cohort* of the present work. Both studies received approval from the McGill Faculty of Medicine Institutional Review Board, and all participants provided written informed consent. The same principal investigator (C.P.) conducted both studies using a similar laboratory protocol in the same experimental setting.

Visit one consisted of screening to collect demographic (all participants) and clinical information (PwP only). No participant had cognitive impairment (Montreal Cognitive Assessment ≥ 26). All participants reported normal or corrected-to-normal vision and the ability to walk independently for 30 minutes. PwP were screened in both OFF and ON dopaminergic medication states. PwP had median (IQR) MDS-UPDRS III scores of 45 (±15) ON and 47 (±10) OFF. All PwP had a clinical diagnosis of idiopathic PD (median ± IQR disease duration: 9 ± 4 years). Exclusion criteria included diabetes, orthopedic disorders, and neurological disorders other than PD. Nine PwP experienced freezing of gait (FoG) (New Freezing-of-Gait Questionnaire part I > 0). All participants were right-hand dominant according to the Edinburgh Handedness Inventory, except for two PwP and one HC who were left-hand dominant. There was no significant difference between PwP and HC in sex (*p* = 0.673, Fisher’s exact test). PwP were older than HC (*p* = 0.005, Mann–Whitney U test).

Visits two and three consisted of two experimental gait tasks (performed OFF for PwP). Visits were separated by at least two days. Participants arrived after a minimum six-hour overnight fast. PwP wore a lower-back inertial sensor (Opal V2R, Clario, Philadelphia, USA; 55 × 40.2 × 12.5 mm; 23.6 g; 128 Hz sampling rate) (Fig. 1). HC walked without sensors and served as neurological controls in this study. All participants received a 185 MBq bolus injection of 18F-fluorodeoxyglucose (18F-FDG) and completed two continuous gait tasks at a self-selected comfortable speed: straight walking (straight condition) and turning (complex condition), in randomized order. Compared to straight walking, complex walking required greater attentional control. Each task began immediately after the 18F-FDG injection. For PwP, inertial recordings covered the first 20 minutes, but walking continued for 30 minutes. HC walked for 40 minutes. These durations satisfied 18F-FDG uptake requirements (∼20 minutes). All participants completed both tasks. FoG episodes were quantified in PwP.

**Fig. 1.**
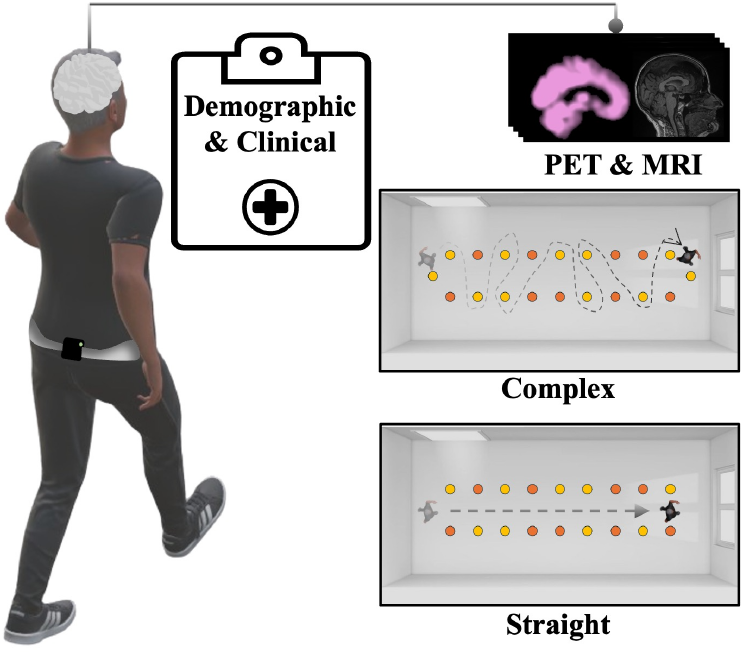
Overview of the *laboratory cohort* protocol (adapted from [22]).

After each gait task, participants underwent 18F-FDG positron emission tomography (PET) and T1-weighted magnetic resonance imaging (MRI) at the Montreal Neurological Institute (MNI). PET acquisition began approximately 50 minutes post-injection. PET preprocessing included co-registration to the individual MRI, spatial normalization to the MNI template, and smoothing with an 8-mm full-width-at-half-maximum Gaussian kernel. Additional details are reported elsewhere [32], [33].

#### 2) Brain Network Extraction

To characterize PD motor network dysfunction during gait, ordinal trend canonical variates analysis (OrT-CVA) was applied. OrT-CVA is a PCA method that identifies covariance networks representing brain regions that co-vary during a task [23]. Each identified network yields an expression score for every participant and condition, quantifying the degree to which the network is expressed. A network is retained when expression scores follow an ordinal trend within participants across conditions, defined as a monotonic increase or decrease between tasks.

Each preprocessed PET image acquired from each participant during each task was restricted to a putative PD motor network using a binary mask. The mask included regions of the PD motor network: putamen, pallidum, thalamus, pons, cerebellum, premotor cortex (PMC), and posterior parietal cortex (PPC) [23]. Each masked image was reshaped into a one-dimensional vector. For each task, participant vectors were assembled into a group-level matrix with 25 rows (participants) and 45,547 columns (voxels of the putative PD motor network). Each matrix was double-centered to generate a subject residual profile matrix (*SRP*_*group*_), and task-independent effects were removed. A subject-by-subject covariance matrix was computed from each *SRP*_*group*_, and eigenvector image networks (*v*_*i*_) were extracted via eigendecomposition. Eigenvectors *v*_*i*_ were scaled by their eigenvalues (*λ*_*i*_) to derive participant-specific unitless expression scores (*u*_*i*_):

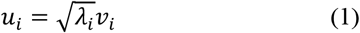

A fixed-effects design matrix encoded group and condition, with straight walking as baseline and complex walking as the condition of interest. The eigenvector *v*_*i*_ that best discriminated between groups and conditions was retained as the final putative PD motor network. This network yielded a unique expression score for each participant and condition, along with its corresponding z-scored voxel map. Voxel contributions within the z-scored map were quantified using bootstrap resampling (1,000 iterations). Positive z-scores indicated relative metabolic increases, and negative z-scores indicated decreases. A region was considered a significant contributor if it contained a cluster with peak |*z*| ≥ 1.64 (∼*p* < 0.05, one-tailed) and a minimum cluster size of 70 voxels.

#### 3) ACI Calculation

ACI, or long-term Lyapunov exponent, is a nonlinear measure of gait automaticity [26], [27]. ACI quantifies deviation from stereotyped locomotor control by capturing increases in dynamic complexity arising from extrinsic (e.g., external cues) or intrinsic (e.g., neural dysfunction) disturbances. This deviation reflects a shift from automatic locomotor control toward less flexible and attention-dependent control. Lower ACI indicates reduced automaticity and greater reliance on attention. Higher ACI indicates greater automaticity and reduced attentional reliance.

ACI was computed in PwP using lower-back inertial sensor acceleration. Briefly, periods disrupting steady-state gait (e.g., long FoG episodes), as well as start and end effects (first and last 20 seconds), were excluded from both tasks. Clean walking segments were linearly detrended, and signals were low-pass filtered using a fourth-order zero-phase Butterworth filter with a 16 Hz cutoff determined by residual analysis. Strides were detected from vertical acceleration [34] and acceleration magnitude (a_mag_) was computed as:

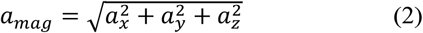

Rosenstein’s algorithm was applied to estimate ACI [35].

Processed *a*_*mag*_ signals were time-normalized to 100 frames per stride to reduce bias arising from differences in walking speed. An embedding dimension of 5 and a time delay of 10 were selected using the global false nearest neighbors and the average mutual information methods, respectively. Divergence was tracked over 4–12 strides. For each task, ACI was computed across the entire 20-minute recording to obtain a single task-specific ACI estimate per participant.

### B. Influence Analysis

#### 1) Overview

Two case studies were conducted to examine whether and how PD motor network dysfunction during gait influences clinical convergence. The first case study used the *laboratory cohort* to evaluate this influence in well-controlled mobility capacity contexts (Fig. 2). The second case study evaluated this influence in a mobility performance context using broader and heterogeneous datasets. In both case studies, motor severity was expressed using MDS-UPDRS III scores converted according to a previously described method [36]: 1 (mild ≤ 32), 2 (32 < moderate ≤ 58), and 3 (severe > 58) for PwP. This conversion aligned with prior work [37] and enabled the integration of ON and OFF MDS-UPDRS III scores into a single score. In contrast to case study 1, HC were included in case study 2 and labeled as 0 for unbiased clinical convergence estimation only [38].

**Fig. 2.**
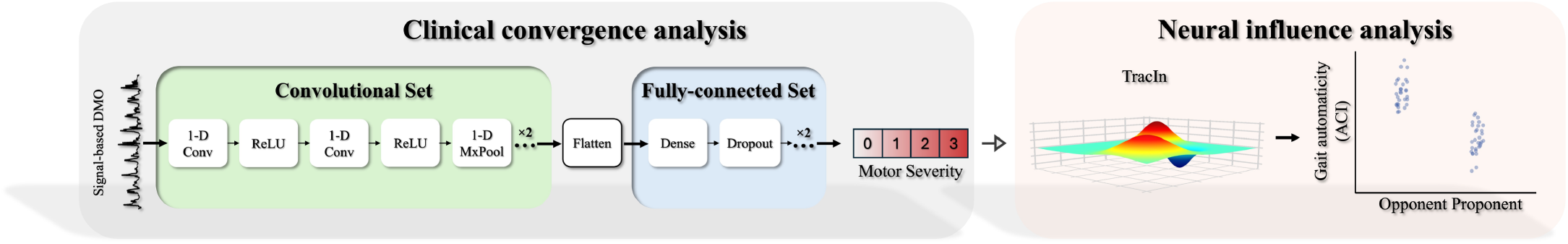
Overview of the experiments conducted in each case study (adapted from [22]).

### 2) Case Study 1

The signal-based DMO corresponded to continuous, non-overlapping 5-second windows of raw lower-back *a*_*mag*_ signals resampled to 100 Hz during both tasks, without the filtering and detrending applied for ACI computation (∼190 valid windows per participant after segmentation) [22], [28]. For each participant, all *a*_*mag*_ windows from both tasks were pooled and labeled with the participant’s motor severity score. Clinical convergence was evaluated using a deep convolutional neural network (CNN) trained to estimate motor severity scores from each 5-second *a*_*mag*_ window. The architecture comprised convolutional layers that extracted deep spatial features followed by fully connected layers that learned the relationships between extracted features and ground-truth severity scores [22], [28]. Nested leave-one-subject-out cross-validation was used to prevent participant-level leakage [39]. At each split, one participant was held out as the independent outer-test set, and the remaining participants formed the outer-training set. Within each outer-training set, an inner loop performed random-search hyperparameter tuning (∼60% of participants for training and 40% for validation). CNN was then retrained on the full outer-training set with the best selected hyperparameters and evaluated on the unseen outer-test set.

Mechanisms influencing clinical convergence were examined using TracIn [31]. Unlike feature-level XAI tools such as saliency maps, TracIn quantifies the influence of individual training observations on specific test set estimations. Influence reflects the extent to which a training observation shifts the model loss for a given test sample during stochastic gradient descent optimization. Training observations that move the prediction toward the correct severity score are called proponents, while those that move the prediction away from the correct score are called opponents. For each test estimation from each participant, TracIn was applied at the best checkpoint [40] to identify the training *a*_*mag*_ windows that acted as proponents or opponents. The average ACI of proponents and opponents was then computed across all predictions for each participant, yielding one aggregated proponent and one aggregated opponent ACI estimate per participant.

#### 3) Case Study 2

Data from three sources were combined to form the *home cohort*. These included the Long-Term Movement Monitoring Database (LTMM) [41], [42], the Levodopa Response Study (LDS) [43], and the daily living dataset from a recent study sponsored by the Michael J. Fox Foundation (MJFF) [44]. LTMM included 66 HC with three days of lower-back inertial sensor data (DynaPort Hybrid, McRoberts, The Hague, Netherlands; 87 × 45 × 14 mm; 74 g; 100 Hz). LDS included 16 PwP with two days of inertial sensor data (Shimmer Research Ltd., Dublin, Ireland; 51 × 34 × 14 mm; 23.6 g; 50 Hz). MJFF included 64 PwP with seven days of inertial sensor data (AX3/6, Axivity Ltd., York, UK; 23 × 32.5 × 7.6 mm; 11 g; 100 Hz). The combined cohort comprised 146 participants: 80 PwP (36% female; median ± IQR age: 66 ± 8 years; MDS-UPDRS III ON: 33 ± 12; OFF: 35 ± 28; disease duration: 10 ± 9 years) and 66 HC (62% female; median ± IQR age: 77 ± 6 years). Additional details are provided elsewhere [41]–[44].

Gait sequences were identified using a validated algorithm commonly applied in PD research [34]. Only sequences from the first two days were retained. Although slightly below current recommendations [45], two days have been shown to be sufficient for reliable capture of real-world gait performance (test–retest reliability > 85% [12]). Furthermore, it reduces potential bias arising from differences in wear time across participants and preserves the heterogeneity required to evaluate the real-world transferability of the findings [38]. ACI was computed for sufficiently long gait sequences using the same procedure as in the *laboratory cohort*, except that sensor orientation was corrected, first and last strides were removed, and low-pass filtering used 13 Hz for LTMM and MJFF and 5 Hz for LDS. Importantly, ACI was computed over 70 strides per sequence, balancing minimal reliability requirements with the shorter walking bouts typical of aging and PD in home settings [46], [47]. Raw *a*_*mag*_ signals from LDS participants were resampled to 100 Hz. All 5-second *a*_*mag*_ windows were then extracted from each detected sequence for all *home cohort* participants (∼900 valid windows per participant after segmentation). CNN evaluated clinical convergence using nested k-fold cross-validation (k = 4) with an inter-subject splitting scheme that ensured homogeneous distribution of severity levels and data sources across folds [39]. TracIn was applied to *home cohort* PwP. Due to computational constraints, TracIn was restricted to 10 randomly selected estimations per PwP. The aggregated ACI of proponents and opponents was computed for each PwP.

### C. Statistics

Normality (Shapiro–Wilk test), equality of variance (Levene’s or Fisher’s test), outliers (Cook’s distance or visual inspection), and multicollinearity (variance inflation factor) were assessed as appropriate for all analyses. *α* was set at 0.05. All *p* values were estimated using one-sided permutation testing (1,000 repetitions) to reduce chance effects and control for type I error. Significance was concluded only when *p* remained below *α* with and without suspected outliers. Effect sizes and 95% confidence intervals (CI) were computed via bootstrapping (1,000 repetitions) and are reported for significant results. Raw statistics are reported.

The presence of an ordinal trend in network expression was assessed within each group (*laboratory cohort*) using permutation testing [23]. To determine whether the extracted network corresponded to the PD motor network: (1) expression scores were compared between PwP and HC within each task using the Mann–Whitney U test and analysis of covariance to examine potential age effects; (2) PwP expression scores were correlated with raw MDS-UPDRS III OFF using Pearson partial correlation controlling for age, sex, and FoG status as covariates of no interest.

To first confirm that ACI reflected reduced automaticity under greater attentional demand, ACI was compared between straight and complex walking in PwP using the Wilcoxon signed-rank test. To determine whether lower ACI related to greater PD motor network dysfunction, ACI and network expression were correlated within PwP for each task using Spearman partial correlation controlling for age, sex, and FoG. To quantify clinical convergence, both case studies used mean absolute error (MAE) and zero-order Spearman rank correlation between participant-level aggregated CNN estimations (mean of all estimations per participant) and ground-truth motor severity scores.

To determine whether and how neural dysfunction influenced clinical convergence, participant aggregated ACI of proponents and opponents were compared using the Wilcoxon signed-rank test. If lower ACI was associated with greater PD motor network dysfunction and (a) was lower in proponents than in opponents, neural dysfunction enhanced convergence; (b) was lower in opponents than in proponents, it impeded convergence; (c) showed no difference, it had no influence.

## III. Results

### A. Quantification of PD Motor Network Dysfunction via ACI

The second eigenvector image was retained as the putative PD motor network. A significant ordinal trend was observed in putative PD motor network expression across tasks in both groups, corresponding to strong suppression of network expression as task complexity increased (Fig. 3(a)). Specifically, 15 of 18 PwP showed a monotonic decrease in network expression across tasks (*p* = 0.002, *r*_*rb*_ = −0.789, *95% CI* [−1.00, −0.45]), as did all HC (*p* = 0.011). Baseline expression during straight walking did not differ between groups (*p* = 0.267). However, task-related suppression during complex walking was strongly reduced in PwP compared to HC (*p* = 0.003, *r*_*rb*_ = −0.746, *95% CI* [−1.00, −0.40]). No age effects were observed on group differences for either straight (*p* = 0.658) or complex walking (*p* = 0.473). The putative PD motor network included clusters of decreased metabolism in the PPC (inferior and superior lobules), PMC, and cerebellum (lobules VIII, VI, and Crus I), and clusters of increased metabolism in the putamen, thalamus, pallidum, cerebellum (lobules IV–V and Crus II), pons, and PPC (angular gyrus) (Fig. 3(b)). Full cluster details are provided in Supplementary Table I, and the z-scored voxel map is available (see support information). Within PwP, putative PD motor network expression during straight walking was not correlated with MDS-UPDRS III OFF scores (Fig. 3(c)). During complex walking, reduced task-related suppression (i.e., higher expression scores) was moderately correlated with MDS-UPDRS III OFF. Task-related suppression of the PD motor network was accompanied by a strong reduction in ACI between straight and complex walking (*p* = 0.023, *r*_*rb*_ = −0.579, *95% CI* [−0.99, −0.11], Fig. 3(d)). Higher network expression during straight walking was moderately correlated with higher ACI, whereas during complex walking, reduced network suppression was moderately correlated with lower ACI (Fig. 3(e)).

**Fig. 3.**
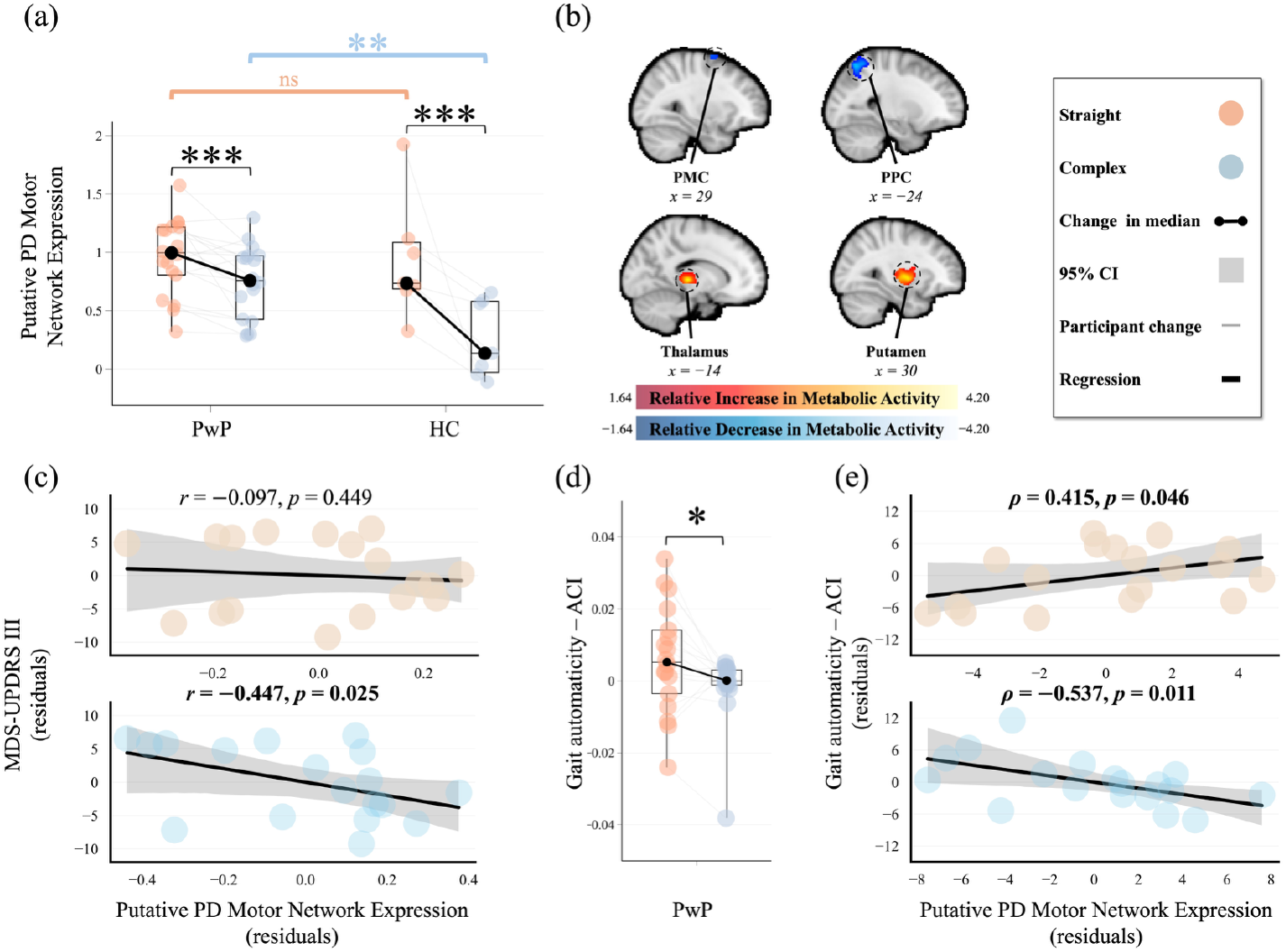
ACI as a measure of PD motor network dysfunction. (a) Box plots of putative PD motor network expression during straight and complex walking by group. (b) Projection of the z-scored voxel-weight map of the putative PD motor network onto the MNI152 template, showing example significant clusters. (c) Pearson partial correlations between raw MDS-UPDRS III scores and network expression during straight and complex walking in PwP. (d) Box plots of gait automaticity (ACI) during straight and complex walking in PwP. ACI values are unitless. (e) Spearman partial correlations between ACI and network expression during straight and complex walking. Bold statistics indicate significant results. No case was overly influential and collinearity remained low. ∗*p* < 0.05, ∗∗*p* < 0.01, ∗∗∗*p* < 0.001, ns indicates non-significant.

### B. Influence on Clinical Convergence

In case study 1 (mobility capacity context), an MAE of 0.127 and a strong correlation were found between CNN estimates and ground-truth motor severity scores (*p* = 0.002, *ρ* = 0.820, *95% CI* [0.55, 0.96], Fig. 4(a)). ACI was strongly reduced in *a*_*mag*_ windows that acted as proponents compared to opponents (*p* = 0.019, *r*_*rb*_ = −0.626, *95% CI* [−1.00, −0.18], Fig. 4(b)). In case study 2 (mobility performance context), an MAE of 0.461 and a strong correlation were found between CNN estimates and ground-truth motor severity scores (*p* < 0.001, *ρ* = 0.813, *95% CI* [0.75, 0.85]). ACI was moderately reduced in *a*_*mag*_ windows that acted as proponents compared to opponents (*p* = 0.023, *r*_*rb*_ = −0.282, *95% CI* [−0.517, −0.037]).

**Fig. 4.**
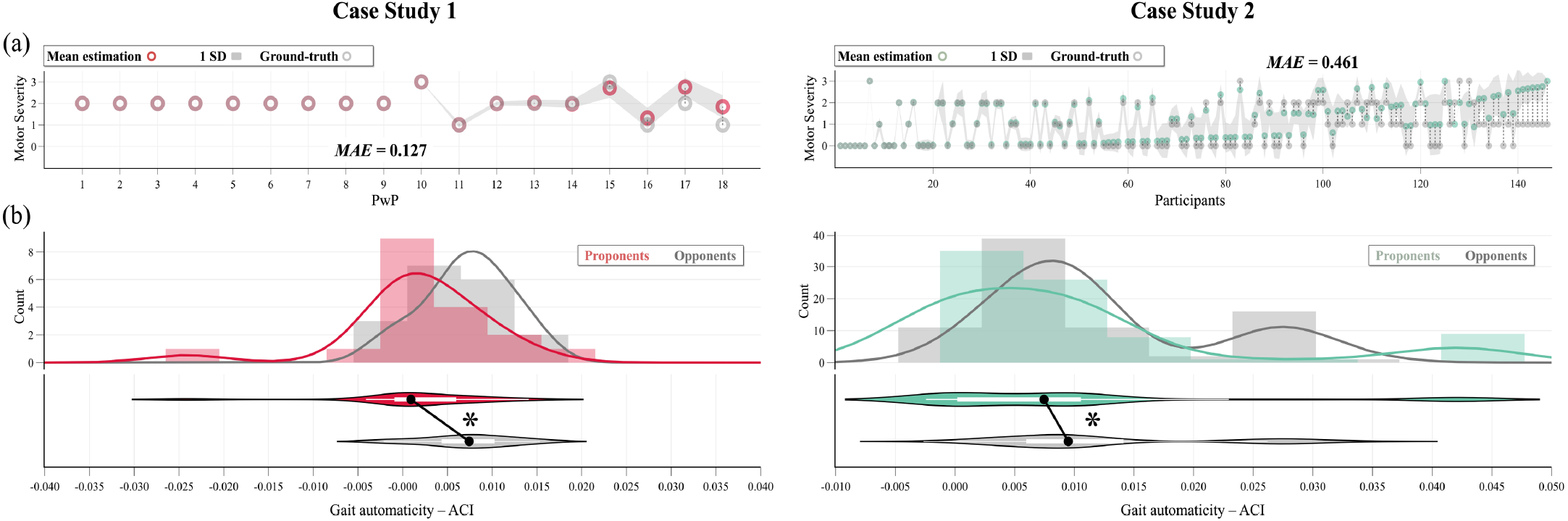
Case studies results. (a) Aggregated estimations and ground-truth motor severity scores (sorted by ascending error), and overall group-level MAE. (b) Distribution plot comparing gait automaticity, expressed by aggregated ACI, between windows acting as proponents and those acting as opponents. ∗*p* < 0.05.

## IV. Discussion

Here, we asked whether ACI can serve as a measure of underlying PD motor network dysfunction and whether this dysfunction drives the convergence of a gait DMO with clinical scales. Two principal findings emerged. First, lower ACI (i.e., altered automaticity) was associated with greater PD motor network dysfunction during gait. Second, lower ACI enhanced the clinical convergence of the gait DMO.

The extracted spatial covariance network exhibited features consistent with the PD motor network described in prior literature, including reduced glucose metabolism in PMC and PPC and increased metabolism in pallidum, putamen, and thalamus [23]. Together with the observed correlation between network expression and motor severity scores, these findings suggest that the identified network represents the true neural correlates of the PD motor network, rather than an unrelated metabolic network.

Several converging results in the present study suggest that ACI reflects PD motor network dysfunction. As walking complexity increased, network expression was suppressed, but this suppression was strongly attenuated in PwP. While baseline network expression during straight walking did not differ between PwP and HC, clear group differences emerged during complex walking. In line with this, network expression within PwP correlated with motor severity during complex walking but not during straight walking, supporting the interpretation that PD motor network dysfunction manifests behaviorally during reduced automatic control. ACI closely mirrored these dynamics and was correlated with network expression, with stronger associations during complex walking, where PD-specific effects were most pronounced. Reduced ACI therefore appears to reflect impaired automatic control linked to PD motor network dysfunction. This mechanistic anchoring may explain why certain gait DMOs, although capturing a single observable feature of mobility (i.e., gait), demonstrate convergence with broad severity scales: they may reflect the expression of a distributed network dysfunction that shapes multiple dimensions of mobility impairment [11], [12].

The DMO consisting of movement magnitude during gait showed strong clinical convergence, with MAE and correlation with ground-truth motor severity scores comparable to or exceeding those reported in similar work [37]. The findings suggest that underlying PD-specific neural dysfunction shapes this clinical convergence. *a*_*mag*_ windows recorded during gait sequences in which ACI was reduced (i.e., more PD motor network dysfunction) exerted a positive influence on severity estimations, whereas windows reflecting preserved automatic control (i.e., less PD motor network dysfunction) exerted a negative influence. This effect was medium to large and persisted across mobility capacity and performance contexts. These results offer a possible mechanistic explanation for prior DMO validation studies reporting increased convergence with clinical scales under walking conditions characterized by reduced automaticity (e.g., complex lab task or home versus straight walking) [11], [12]. When automatic control is taxed, PD motor network dysfunction may become more salient, gait DMOs may more strongly capture underlying neural dysfunction, and clinical convergence may increase.

Several limitations warrant consideration. First, the laboratory protocol required prolonged walking, and ACI required a high number of strides. While longer gait sequences provide more informative representations of mobility [19], future work should investigate measures computable on shorter sequences because real-world walking bouts in PwP are typically shorter [46], particularly in individuals with more deteriorated gait patterns such as PwP with FoG. In our *laboratory cohort*, PwP with FoG showed particularly low ACI values (often < 0 in both tasks) and did not systematically decrease ACI in response to increases in task complexity (Fig. 3(c)). Although this is consistent with the known reduced flexibility observed in FoG [24], such low ACI values may also reflect subthreshold disturbances despite careful processing, such as micro-FoG episodes, which can alter complexity estimates without necessarily reflecting reduced automaticity. Second, the sample size of the *laboratory cohort* was modest and included relatively well-functioning individuals, limiting the generalizability of the findings. Third, the design was cross-sectional, and only one gait DMO was evaluated, whereas other DMOs may show different mechanistic relationships [48]. Fourth, influence was treated categorically rather than continuously (i.e., whether a window enhanced or reduced clinical convergence without quantifying the magnitude of its influence). Finally, mobility perception was not considered as an additional construct [17], [19].

## V. Conclusion

In conclusion, the findings suggest that altered gait automaticity indexed by ACI reflects PD motor network dysfunction and that this dysfunction drives the convergence of gait a DMO with clinical scales.

This study is the first to apply OrT-CVA to task-based gait PET data, showing that a participant-level multivariate approach can link brain network dynamics to gait DMOs in PD. In parallel, altered automaticity quantified via ACI emerged as a promising gait-based proxy of neural dysfunction. The application of TracIn in PD DMO research further strengthens the contribution. PD is a complex disorder, and big data approaches are often proposed to address this complexity. However, the use of advanced analytical approaches such as deep learning remains limited by concerns regarding explainability [49]. Here, we show that a modern XAI tool can provide mechanistic insight into the relationships between digital and clinical measures and potentially reduce reluctance toward AI-based methods in PD DMO research. Evaluation in both mobility capacity and performance contexts enhances the transferability of the findings.

Beyond methodological advances, the clinical implication is central. Gait DMOs hold potential for disease-modifying trials, where they may serve as exploratory outcomes [50], as well as for clinical practice by enabling granular mobility monitoring [8], [9]. Convergent validity is a core component of construct validity, which conditions regulatory approval and clinical adoption of gait DMOs [17]. Multilevel validation frameworks are increasingly viewed as necessary, and the integration of mechanistic evidence appears relevant within this context. By showing that neural dysfunction shapes clinical convergence, this work supports integrating mechanistic evidence to enhance the convergent (and construct) validity of gait DMOs. Future work should extend this approach beyond a single signal-based DMO to other gait DMOs [8]–[10], broader mobility domains [48], and composite DMOs [8].

## Data Availability

Codes available at https://github.com/cyrillemvomo/PD_Gait_DMO_XAI, and the brain map at https://neurovault.org/collections/MFZKQGET/.

## Notes

This work was supported in part by Parkinson Canada, the Natural Sciences and Engineering Research Council of Canada, and the Quebec BioImaging Network.

### Competing Interest Statement

The authors have declared no competing interest.

### Author Declarations

This study was approved by the McGill Faculty of Medicine Institutional Review Board for Human Subjects and all participants provided written informed consent in accordance with the Declaration of Helsinki before entering the study.

