## Supplementary Table 1 for "Gait-Related Digital Mobility Outcomes in Parkinson’s Disease: New Insights into Convergent Validity?"

**Supplementary Table 1 – Relative increased and decreased metabolism**

| Peak *z*-score | x | y | z | Peak z location (AAL) |
|---|---|---|---|---|
| 3.96 | 30 | -4 | 0 | Putamen_R |
| 3.92 | -14 | -24 | 2 | Thalamus_L |
| 3.81 | 48 | -50 | 34 | Angular_R |
| 3.55 | 12 | -48 | -24 | Cerebelum_4_5_R |
| 3.07 | -12 | 4 | -6 | Pallidum_L |
| 3.02 | 8 | -20 | 18 | Thalamus_R |
| 2.58 | 44 | -38 | -42 | Cerebelum_Crus2_R |
| -4.04 | -28 | -40 | -48 | Cerebelum_8_L |
| -3.53 | -24 | -66 | 56 | Parietal_Sup_L |
| -3.31 | 14 | -64 | 60 | Precuneus_R |
| -3.3 | 42 | -74 | -30 | Cerebelum_Crus1_R |
| -2.8 | 58 | -42 | 48 | Parietal_Inf_R |
| -2.73 | 34 | -38 | -32 | Cerebelum_6_R |
| -2.48 | 28 | -2 | 70 | Frontal_Sup_R |
| -2.29 | -46 | -34 | 38 | Parietal_Inf_L |

Coordinates are given in MNI space. AAL corresponds to the Automated Atlas Labeling 3.1.
